# Frequency of nursing student medication errors: A systematic review

**DOI:** 10.1101/2023.02.26.23286460

**Authors:** Christos Triantafyllou, Maria Gamvrouli, Pavlos Myrianthefs

## Abstract

**Introduction:** Health promotion and patient safety are the main targets of the healthcare provision by the National Health Systems. As for the nursing profession, nursing students make medication errors during clinical interventions, which could be a potential danger to patient safety.

**Aim:** The investigation of the frequency of nursing student medication errors, as well as the frequency of each type of nursing student medication errors.

**Methods:** A systematic review of the literature was conducted on the electronic database “PubMed” with the keywords: “medication error”, “prescribing error”, “drug error”, “drug use error”, “drug mistake”, “wrong drug”, “wrong dose”, “administration error”, “dispensing error”, “incorrect drug”, “incorrect dose”, “inappropriate prescribing”, “inappropriate medication”, “transcription error”, “nursing student”, “nursing trainee” and on the Greek electronic database IATROTEK-online with the keywords: “medication errors” and “nursing students”, without time limit for the publication of scientific papers. On PubMed, the keywords were searched in the title and abstract of the studies. Studies were excluded if they were not published in English and Greek language, were conducted on animals, and were case studies, editorials, and letters to the editor.

**Results:** Of the 47 scientific papers retrieved, 6 were included in the systematic review. A total of 1,904 nursing student medication errors were recorded by nursing students. The majority of errors were: 1) wrong dose form (330,17%), 2) omission error (313, 16.4%), and 3) wrong time (259, 13.6%).

**Conclusions:** The frequency of nursing student medication errors is high. The safe administration of medications is an important skill that nursing students should learn. At a theoretical and practical/clinical level, it would be advisable for clinical nurses and academics to jointly develop an educational program to acquire correct knowledge and perceptions regarding safe medication administration.

## INTRODUCTION

Patients’ safety is one of the key objectives of National Health Systems, which can be affected by various factors. One of the most important factors affecting patient safety is medication errors made by healthcare professionals.^1^ In America, medication errors are the third leading cause of death, causing 50,000 to 100,000 deaths per year.^2^ In hospitals, medication errors, urinary tract infections associated with urinary catheters, falls, and immobilization are the most common causes of patient injury.^3^ Many studies have suggested that medication errors cause about one-third of complications for hospitalized patients.^4,5^

The reasons for medication errors can range from lack of knowledge to increased workload to non-compliance with guidelines. The mistakes can occur at many stages of the treatment process, including prescribing, preparing, dispensing, and administering medications. Medication administration errors account for most errors, however.^6^ Nurses commit medication errors more frequently than physicians in everyday clinical practice, although medication errors can be caused by a wide array of healthcare professionals.^7,8^ During their clinical practice, nursing students are more likely to take risks and make errors than professional nurses because of their underdeveloped skills, limited clinical experience, and lack of knowledge.^9^ There have been various reports describing different types of medication errors by nursing students, which have led to complications of varying severity.^10^

During nursing students’ studies, Sulosaari argues that clinical skills are not adequately taught and, therefore, they need to receive a more comprehensive education in applying clinical skills, especially in preparing and administering medications.^11^

This systematic review aimed to investigate the frequency of nursing student medication errors and the frequency of each type of nursing student medication errors.

## METHODS

We conducted a systematic literature review utilizing the international electronic databases PubMed and IATROTEK-online to investigate the frequency and type of nursing student medication errors.

The following keyword combination was used for the PubMed search strategy: ((((((((((((((“medication error”[Title/Abstract]) OR (prescribing error*[Title/Abstract])) OR (drug error*[Title/Abstract])) OR (Drug Use Error*[Title/Abstract])) OR (drug error*[Title/Abstract])) OR (wrong drug*[Title/Abstract])) OR (wrong dose[Title/Abstract]))) OR (administration error*[Title/Abstract])) OR (dispensing error*[Title/Abstract])) OR (incorrect drug*[Title/Abstract])) OR (incorrect dose[Title/Abstract])) OR (inappropriate prescribing [Title/Abstract])) OR (inappropriate medication [Title/Abstract])) OR (transcription error*[Title/Abstract])) AND ((nursing student*[Title/Abstract]) OR (nursing trainee*[Title/Abstract])), without a time limit regarding the publication date of the studies. In IATROTEK-online, the search strategy was as follows: medication errors, nursing students, without a time limit regarding the publication date of the studies.

This systematic review included studies that met the following criteria:

- Studies were conducted on humans.
- Studies focused on medication errors made by nursing students.
- Studies included data concerning the frequency of each type of medication error made by nursing students.
- Studies were published in Greek or English.
- On the other hand, the literature exclusion criteria were as follows:
- Studies were case studies, editorials, and letters to the editor.
- Studies were not in English and Greek language.
- Studies were conducted on animals.

The following data were extracted from each study after the selection of the studies included in the systematic review: first author’s name, the country of origin of the study, the publication year, the type of study, the purpose of the study, the sample studied, the frequency of medication errors made by nursing students.

## RESULTS

### Characteristics of Systematic Review Studies

There were 47 articles reviewed, 29 of which were rejected due to a non-relevant title or abstract. In the end, based on the inclusion and exclusion criteria, six studies were included in this systematic review after reading the full text of 18 articles (Figure 1)^12–17^

**Figure 1:**
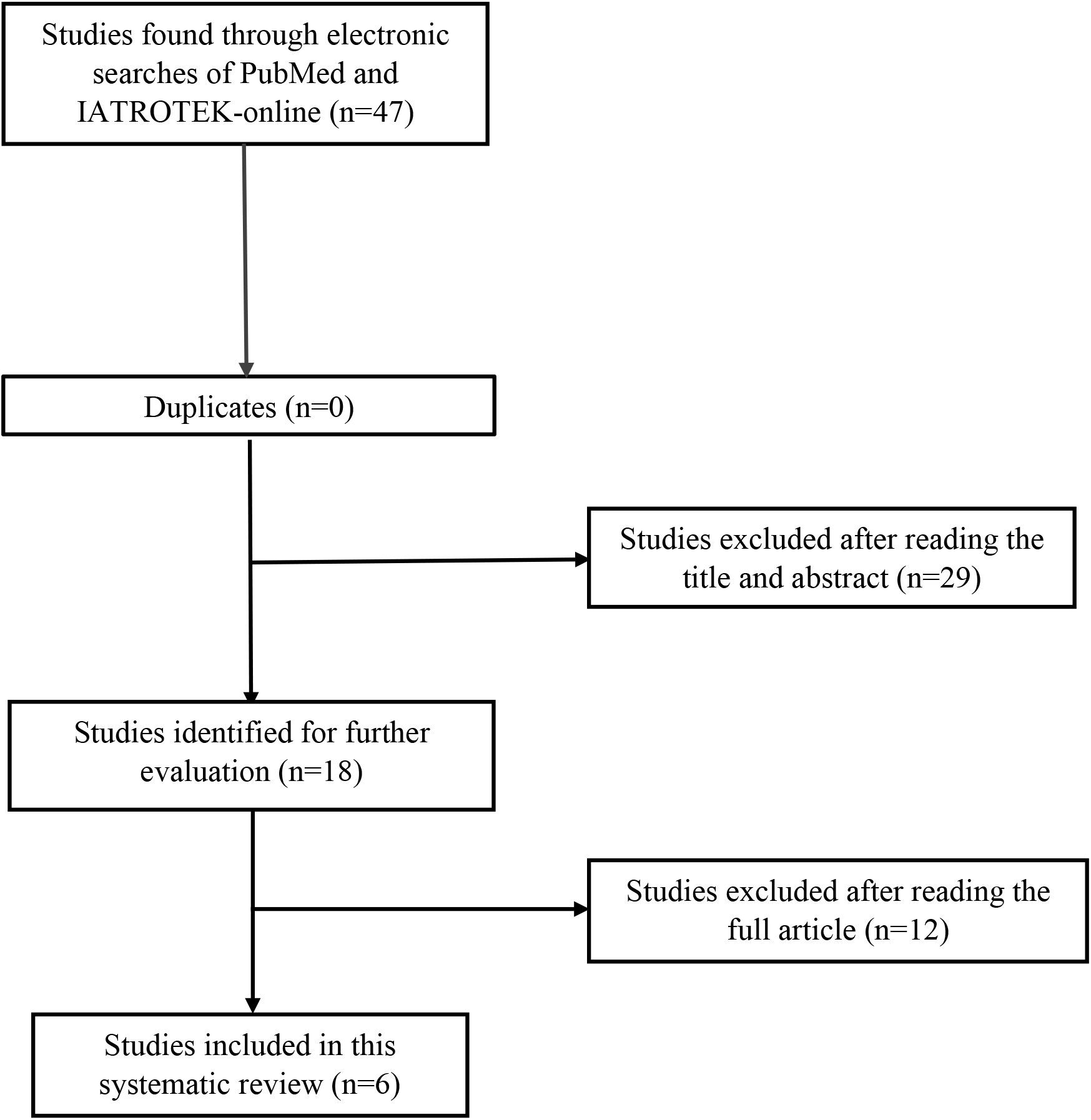
Flow chart of the systematic review

A summary of the study characteristics is presented in **Table 1**. The majority of the studies were conducted in the American continent, specifically, two in the USA^16,17^, and two in Canada^13,15^. The remaining studies were conducted in Asia, specifically, one in Turkey^12^ and one in the Philippines.^14^ In terms of study design, the majority of the studies were cross-sectional studies^12,14,15^ two were retrospective observational studies ^13,16^ and one was descriptive study.^17^

**Table 1.** Summary of the studies in the systematic review

| **First author,**<br>**Study design, Country** | **Purpose of the study** | **Study sample** | **Frequency of the most common medication errors** |
| --- | --- | --- | --- |
| Valdez et al<sup>14</sup>,<br>Cross-sectional study,<br>Philippines | Investigating the factors leading to medication errors by nursing students | 329 nursing students | Total errors=62<br>Wrong time= 25 (40%)<br>Missed dose = 26 (42%)<br>Wrong dose=10 (16%) |
| Wolf et al<sup>17</sup>,<br>Descriptive study,<br>USA | The evaluation of medication error reports conducted by nursing students | 27 reports of medication errors | Total errors=27<br>Incorrect administration technique=7 (26%)<br>Missed dose=7 (26%)<br>Wrong dose =6 (22%)<br>Wrong time=2 (7.4%) |
| Wolf et al<sup>16</sup>,<br>Retrospective observational studies,<br>USA | To investigate the characteristics of medication errors conducted by nursing students | 1208 reports of medication errors | Total errors=1208<br>Missed dose = 248 (19%)<br>Wrong dose =224 (17.1%)<br>Wrong time= 221 (17%) |
| Harding and Petrick<sup>13</sup>,<br>Retrospective observational study,<br>Canada | To investigate the characteristics of medication errors conducted by nursing students | 173 reports of medication errors | Total errors = 173<br>Missed dose=32 (18,5%) |
| Cebeci et al<sup>12</sup>,<br>Cross-sectional study,<br>Turkey | The frequency and type of medication errors made by nursing students | 324 nursing students | Total errors=402<br>Non-compliance with the aseptic technique = 96 (23.8)<br>Wrong dose = 90 (22.8%)<br>Failure to report given medicines = 73 (18.1%)<br>Wrong time=5 (1.2%) |
| Walsh et al<sup>15</sup>,<br>Cross-sectional study,<br>Canada | Factors that increase the likelihood of reporting medication errors made by nursing students | 106 nursing students | Total errors=31<br>Failure to report the medicine given = 6 (18%)<br>Wrong time=6 (18%) |

### Frequency of medication errors by nursing students

According to Table 1, nursing students made 1904 medication errors. According to Table 1, nursing students made 1904 medication errors. A large majority of errors involved administering the wrong dose (330, 17%), omitting a dose (313, 16.4%), or administering the medication at the wrong time (259, 13.6%) (Table 1).

## DISCUSSION

In this systematic review, as previously reported, the purpose was to determine the frequency of nursing medication errors made by nursing students, as well as the incidence of different types of nursing medication errors. As far as we know, this is the first systematic review in Greek and international literature that contains studies that have not been conducted in only one country, as in Dehvan et al.’s systematic review.^10^

Based on the findings of the six studies, nursing students are often found to make medication errors. As the medication administration process is not closely monitored and there is no weighted system for properly recording and reporting medication errors, it is expected that the prevalence of medication errors carried out by nursing students is higher than reported in the studies.^18^ There are also some medication errors which remain unreported due to the negative reactions of clinical trainers and nursing management.^18^ Even though the cause of medication errors by nursing students is still unknown, learning the “5 rights” rule regarding the administration of medications, namely, “right patient”, “right drug”, “right dose”, “right route of administration”, and “right time”, will greatly reduce the likelihood of medication errors.^19^ Although nursing students work diligently to assist the therapeutic team in providing care to the patient, they often make mistakes due to a lack of clinical knowledge or clinical experience, which usually go unreported.

A significant finding of this study is that nursing students most commonly made medication errors by administering the wrong dose of the medication. This finding is in accordance with a previous systematic review published in 2021 that included studies conducted only in Iran.^10^

This systematic review has several limitations that should be mentioned. A key finding of the review is the lack of published studies from European and Latin American countries, which makes it difficult to draw firm conclusions about the frequency of medication errors among nursing students, as well as regional variations. Moreover, we acknowledge the possibility of language bias due to the fact that only English-language studies were included in this systematic review. Practical reasons, mainly the difficulty of translating from a variety of languages, led us to the decision to exclude studies published in a different language than English. In addition, searching bibliographies only in international electronic databases may have introduced publication bias to our systematic review because it is unlikely to detect studies that have not been published in peer-reviewed journals. Additionally, potentially related databases like EMBASE and Cinahl were not used.

In conclusion, the frequency of nursing medication errors among nursing students is high. It is important for nursing students to learn how to administer medications safely. Clinical nurses and academics should collaborate to develop an educational program regarding safe medication administration at both theoretical and practical/clinical levels.

## Data Availability

All data produced in the present study are available upon reasonable request to the authors.

## BIBLIOGRAPHY

1. Dimant J. Medication errors and adverse drug events in nursing homes: problems, causes, regulations, and proposed solutions. J Am Med Dir Assoc 2001;2(2):81–93.

2. Seiden SC, Galvan C, Lamm R. Role of medical students in preventing patient harm and enhancing patient safety. Qual Saf Health Care 2006;15(4):272–276.

3. Asensi-Vicente J, Jiménez-Ruiz I, Vizcaya-Moreno MF. Medication Errors Involving Nursing Students: a Systematic Review. Nurse Educ 2018;43(5):E1–5.

4. Kaushal R, Bates DW, Landrigan C, McKenna KJ, Clapp MD, Federico F, et al. Medication errors and adverse drug events in pediatric inpatients. JAMA 2001 25;285(16):2114–2120.

5. Stratton KM, Blegen MA, Pepper G, Vaughn T. Reporting of medication errors by pediatric nurses. J Pediatr Nurs 2004;19(6):385–392.

6. Farzi S, Abedi H, Ghodousi A, Yazdannik A. Medication Errors Experiences of Nurses who Working in Hospitals of Isfahan at 1391. J Qual Res Health Sci 2014;2(4):310–319.

7. Fathi A, Hajizadeh M, Moradi K, Zandian H, Dezhkameh M, Kazemzadeh S, et al. Medication errors among nurses in teaching hospitals in the west of Iran: what we need to know about prevalence, types, and barriers to reporting. Epidemiol Health 2017;39:e2017022.

8. Jember A, Hailu M, Messele A, Demeke T, Hassen M. Proportion of medication error reporting and associated factors among nurses: a cross sectional study. BMC Nurs 2018;17:9.

9. Dehvan F, Mokhtari Z, Aslani M, Ebtekar F, Gheshlagh RG. The prevalence of needlestick injury in students of medical sciences universities: a systematic review and meta-analysis. Hayat 2018;24(2):140–151.

10. Dehvan F, Dehkordi AH, Gheshlagh RG, Kurdi A. The Prevalence of Medication Errors Among Nursing Students: a Systematic and Meta-analysis Study. Int J Prev Med 2021;12:21.

11. Mrayyan MT, Shishani K, Al-Faouri I. Rate, causes and reporting of medication errors in Jordan: nurses’ perspectives. J Nurs Manag 2007;15(6):659–670.

12. Cebeci F, Karazeybek E, Sucu G, Kahveci R. Nursing students’ medication errors and their opinions on the reasons of errors: a cross-sectional survey. J Pak Med Assoc 2015;65(5):457–462.

13. Harding L, Petrick T. Nursing student medication errors: a retrospective review. J Nurs Educ 2008;47(1):43–47.

14. Valdez LP, de Guzman A, Escolar-Chua R. A structural equation modeling of the factors affecting student nurses’ medication errors. Nurse Educ Today 2013;33(3):222–228.

15. Walsh LJ, Anstey AJ, Tracey AM. Student perceptions of faculty feedback following medication errors - A descriptive study. Nurse Educ Pract 2018;33:10–16.

16. Wolf ZR, Hicks R, Serembus JF. Characteristics of medication errors made by students during the administration phase: a descriptive study. J Prof Nurs 2006;22(1):39–51.

17. Wolf ZR, Hicks RW, Altmiller G, Bicknell P. Nursing student medication errors involving tubing and catheters: a descriptive study. Nurse Educ Today 2009;29(6):681–688.

18. Mohammad Nejad I, Hojjati H, Sharifniya SH, Ehsani SR. Evaluation of medication error in nursing students in four teaching hospitals in Tehran. Iranian Journal of Medical Ethics and History of Medicine 2010;3(1):60–69.

19. Dolansky MA, Druschel K, Helba M, Courtney K. Nursing student medication errors: a case study using root cause analysis. J Prof Nurs 2013;29(2):102–108.

